# Impact of Proactive Integrated Care on Chronic Obstructive Pulmonary Disease

**DOI:** 10.1101/2020.05.19.20107110

**Authors:** Patricia B. Koff, Sung-joon Min, Debora L. P. Diaz, Tammie J. Freitag, Shannon S. James, Norbert F. Voelkel, Derek J. Linderman, Fernando Diaz del Valle, Richard K. Albert, Todd M. Bull, Arne Beck, Thomas J. Stelzner, Debra P. Ritzwoller, Christine M. Kveton, Stephanie Carwin, Moumita Ghosh, Robert L. Keith, John M. Westfall, R. William Vandivier

## Abstract

**Background**. Up to 50% of COPD patients do not receive recommended care for COPD. To address this important issue, we developed Proactive Integrated Care (Proactive iCare), a healthcare delivery model that couples integrated care with remote monitoring. **Methods**. We conducted a prospective, quasi-randomized clinical trial in 511 patients with advanced COPD, or a recent COPD exacerbation, to test whether Proactive iCare impacts patient-centered outcomes and healthcare utilization. Patients were allocated to Proactive iCare (n = 352) or Usual Care (n = 159), and were examined for changes in quality of life using the St. George’s Respiratory Questionnaire (SGRQ), symptoms, guideline-based care, and healthcare utilization. **Findings**. Proactive iCare improved the total SGRQ by 7 – 9 units (p < 0.0001), symptom SGRQ by 9 units (p < 0.0001), activity SGRQ by 6 – 7 units (p < 0.001) and impact SGRQ by 7 – 11 units (p < 0.0001) at 3, 6 and 9 months, compared with Usual Care. Proactive iCare increased the 6-minute walk distance by 40 m (p < 0.001), reduced COPD-related urgent office visits by 76 visits per 100 subjects (p < 0.0001), identified unreported exacerbations, and decreased smoking (p = 0.01). Proactive iCare also improved cough, sputum, shortness of breath, the BODE index and oxygen titration (p < 0.05). Mortality in the Proactive iCare group (1.1%) was not significantly different than mortality in the Usual Care group (3.8%; p = 0.08). **Interpretation**. Results suggest that linking integrated care with remote monitoring improves the lives of people with advanced COPD.

## Introduction

In the United States, chronic obstructive pulmonary disease (COPD) affects over 16 million people, costs $32 billion per year, and is the fourth leading cause of death^1-4^. Exacerbatins of COPD are among the most devastating complications of COPD, because they greatly increase the risk of death and account for 60% of COPD costs ^3^ Efforts have focused on standardizing recommendations for therapies that improve symptoms and limit exacerbations, identifying patients who might benefit from these treatments, and increasing compliance with recommended therapies ^1,3^ Surprisingly, only ~50% of COPD patients receive recommended therapies for chronic COPD and COPD exacerbations ^5-7^, up to two thirds of COPD exacerbations are not reported to healthcare providers ^8-10^, and only half of primary care physicians use COPD guidelines ^11-14^ Therefore, novel methods to improve healthcare delivery could bring substantial benefits.

Integrated care is a model of healthcare designed to improve delivery of care by increasing access to care and providing disease and self-care focused education. Enthusiasm for integrated care in patients with COPD was high after several studies showed that it decreased symptoms, improved healthcare delivery, enhanced quality of life and reduced healthcare utilization ^15-19^. But this excitement dampened after two well-designed clinical trials ^20,21^ found that integrated care was expectedly associated with increased COPD-related hospitalizations, emergency department visits and all cause mortality for reasons that remain unclear ^22-24^.

Proactive iCare is a healthcare delivery model that couples traditional integrated care with remote monitoring by healthcare providers to enhance communication, education, recognition of exacerbations, and delivery of guideline-based care ^25,26^. It is designed to identify and solve problems early, because it does not solely depend on the ability of patients to recognize and act on chronic or impending health problems. We performed two studies of Proactive iCare in patients with advanced COPD ^25,26^. The first study randomized 40 COPD patients to Proactive iCare or Usual Care for 3 months, and showed that Proactive iCare improved quality of life and identified unreported exacerbations ^25^ The second study used a pre-post design to examine the impact of Proactive iCare on 100 rural COPD patients over a 3-month period, and showed that Proactive iCare improved quality of life, guideline-based care, recognition of unreported exacerbations and COPD-related health care utilization ^26^. The current study was designed to test the ability of Proactive iCare to improve patient-centered outcomes and healthcare utilization in a larger group of patients with advanced COPD from multiple sites over a 9-month period.

## MATERIALS AND METHODS

### Study Design

We conducted a prospective, quasi-randomized clinical trial to determine the effect of Proactive iCare on patients with advanced COPD over a 22-month period between September 2006 and June 2008. The funding organization would not allow a traditional randomized, controlled clinical trial design for legislative reasons (*i.e*., Colorado Amendment 35), so a quasi-randomized design was used instead. The Colorado Multiple Institutional Review Board approved the study and all subjects completed written, informed consent. This trial was registered at www.clinicaltrials.gov (NCT01044927).

#### Participants and Study Setting

Subjects were recruited from primary care and pulmonary specialty clinics at University of Colorado Hospital, Kaiser Permanente Colorado, Denver Veterans Affairs Medical Center and primary care practices within the Colorado front-range urban corridor. Patients were also recruited from rural counties in part through collaboration with the High Plains Research Network.

Inclusion criteria required the patients to have 1) severe to very severe COPD^27^ or 2) COPD with a FEV_1_ ≥ 50% predicted and a COPD exacerbation within the past year, 3) home access to a standard land telephone line, and 4) legal US and Colorado residency. Subjects were excluded for 1) a clinical diagnosis of asthma, 2) co-existing medical conditions that were likely to cause death within two years, 3) roentgenographic evidence of interstitial lung disease or other non-COPD related pulmonary diagnoses at the time of enrollment, 4) participation in another treatment study, 5) inability or unwillingness to cooperate with self-monitoring and reporting components, 6) inability to perform pulmonary function testing or a six-minute walk, 7) prisoners, pregnant women, or institutionalized patients, 8) current alcohol or drug abuse, 9) non-English speakers, or 10) inability to complete consent.

### Interventions

#### Disease Management

Proactive iCare consisted of a 9-month disease management program that included 1) COPD education; 2) exacerbation education; 3) direct communication with study coordinators; and 4) remote home monitoring^25^. The monitoring equipment included a telecommunication platform (Bosch Health Buddy®, Robert Bosch Healthcare, Palo Alto, CA), a finger pulse oximeter (Onyx II Finger Pulse Oximeter 9550, Nonin Medical, Plymouth, MN), a hand-held spirometer (Microlife #PF 100, Dunedin, FL), and a pedometer (Health Measures SXC America on the Move, Stillwater, MN).

COPD education was given to each subject during the 2 hour enrollment session, during weekday sessions with the Health Buddy®, and informally during phone calls with study coordinators^27^. Subjects were taught to recognize COPD exacerbations, the importance of early therapy with quick-relief medications, and the use of oral corticosteroids and/or antibiotics when needed. Patients were encouraged to call their healthcare provider anytime or study coordinator (9AM to 5PM Monday through Friday) if they suspected the onset of a COPD exacerbation.

Each weekday, subjects would participate in a Health Buddy® session at home lasting approximately 20 minutes. During this session they would receive COPD education^27^ and answer symptom-based questions. They would measure their FEV_1_ and SpO_2_ at rest. Subjects would then walk for 6 minutes and measure the distance walked via a pedometer and the post-exertion SpO_2_. This information was entered into the Health Buddy, analyzed by predetermined algorithms, and categorized into three color-coded groups: green for stable, yellow advising caution and red indicating a possible decline in health status (Table 1). Data were transferred to a database overnight for coordinators to view the next weekday. Coordinators called all patients with red flags and used discretion in contacting patients who had persistent red or yellow flags. Coordinators helped resolve clinical problems directly or by calling the subject’s primary care provider.

**Table 1.** Flag Definitions.

| **Flags** | **Green Flag** | **Yellow Flag** | **Red Flag** |
| --- | --- | --- | --- |
| Shortness of breath | None | With activity | At rest |
| Cough | None/Usual | More than usual |  |
| Sputum | Usual/White | Yellow | Green/Red/Brown |
| Fever | $\leq 101.5^{\circ}\text{F}$ | | $>101.5^{\circ}\text{F}$ |
| Wheeze in past 48 hours | No | Yes |  |
| Activity difficulties | None | Some | Yes |
| FEV<sub>1</sub>, % below baseline | 0-14 | 15-24 | $\geq 25$ |
| 6MWD | Walked | No walk $\leq 1$ day | No walk $> 1$ day |
| Resting SpO<sub>2</sub>, % | $\geq 90$ | 88-89 | $\leq 87$ |
| Post-exercise SpO<sub>2</sub>, % | $\geq 90$ | 88-89 | $\leq 87$ |

*Definition of abbreviations:* 6MWD = 6 minute walk distance, SpO<sub>2</sub> = pulse oximetry oxygen saturation.

Study coordinators called subjects in the Proactive iCare Group at 3 and 6 months to administer the St. George’s Respiratory Questionnaire (SGRQ) and collect data on healthcare utilization. Coordinators met with subjects in person at 9 months to collect selfreported healthcare utilization, administer questionnaires, perform lung function testing and a six minute walk test (6MWT), and to retrieve equipment.

#### Usual Care

Subjects enrolled into the Usual Care group continued to receive care by their healthcare provider. They did not receive COPD education or advice, but were advised to inform their healthcare provider if they had a resting oxygen saturation ≤ 88% at enrollment.

Study coordinators called Usual Care subjects at 3 and 6 months to administer the SGRQ and collect data on healthcare utilization. Coordinators met with subjects at 9 months in person to collect self-reported healthcare utilization, administer questionnaires, perform lung function testing and a 6MWT.

#### Outcomes

The primary outcome was healthcare costs with data coming from University of Colorado and Kaiser Permanente. Quality of life was a key secondary outcome measured by the SGRQ at baseline, 3, 6 and 9 months. Other secondary outcomes were measured at baseline and 9 months. These included respiratory symptoms (*i.e*. cough, sputum production and shortness of breath measured by the modified Medical Research Council [mMRC] Dyspnea scale), COPD and non-COPD-related health care utilization and the 6MWT. We also assessed use of guideline-based care and the number of COPD exacerbations. Self-reported COPD and Non-COPD healthcare utilization was assessed for the previous 12 months at enrollment and compared to healthcare utilization at the study conclusion. A COPD exacerbation was defined as a worsening of a subject’s respiratory signs or symptoms that lead to a change in treatment, including quick-acting medications, and oral corticosteroids or antibiotics.

#### Randomization and Masking

Subjects were allocated to experimental groups using a continuously rotating enrollment schedule with four-day enrollment blocks. Subjects were enrolled into the Proactive iCare group during the first three days of the block, and into the Usual Care group during the last day.

### Sample Size

The study was originally powered to determine the effect of Proactive iCare on healthcare costs in urban patients with COPD, and quality of life in urban and rural patients compared to usual care after one year. The power calculation was based on a prior small study of Proactive iCare *versus* Usual Care^25^. In this study Proactive iCare reduced healthcare costs by $1,401 ± $10,717, compared with an increase of $1,709 ± 12,570 in the Usual Care group^25^. Assuming the same mean difference and standard deviation with a 2:1 allocation, a sample size of 243 in the Proactive iCare group and 121 in Usual Care group would result in an 82% power to detect a difference using a 2-sided t-test with a Type I error rate = 0.05. The study was also powered to determine the effect of Proactive iCare on quality of life measured by the SGRQ in the subgroup analyses. Prior work suggested that Proactive iCare improved the total SGRQ by 10.3 ± 14.9 units compared to 0.6 ±12.3 in the Usual Care group^25^. A sample size of 81 rural subjects in Proactive iCare group and 40 in Usual Care group would yield a 96% power to detect a difference using a 2-sided t-test with a Type I error rate = 0.05. Allowing for 10% attrition, recruitment of 400 intervention and 200 control subjects would be sufficient to test the main hypotheses.

### Statistical Methods

Statistical analyses were performed using SAS for Windows, version 9.2 (SAS Institute, Cary, NC). Comparisons between study participants in the Proactive iCare and Usual Care groups were made (using t-tests or Wilcoxon tests if the distributions were skewed) for continuous variables and chi-square tests (or Fisher’s exact tests if cell sizes were small) for categorical variables. All tests were 2-sided with a significance level of P < 0.05. Changes from baseline in quality of life were determined for the SGRQ at 3, 6 and 9 months for the Proactive iCare and Usual Care groups. Differences between groups were then tested using t-tests. Linear mixed effects models for total SGRQ score including a quadratic time term were considered as an intention-to-treat analysis utilizing participants with incomplete data, and compared with the analyses of participants with complete data.

Symptoms, guideline-based care, and 6MWT distances were compared between baseline and 9 months, and differences were tested using t-tests for continuous variables, and logistic regression including group by time interaction with Generalized Estimating Equations (GEE) to account for the intra-class correlations arising from paired measurements for each patient for dichotomous variables (except chi-square tests when only 9-month measurements were considered). Self-reported COPD and Non-COPD healthcare utilization was assessed for the previous 12 months at enrollment and compared to healthcare utilization at the study conclusion. Differences in 9-month rate of healthcare utilization were generated and compared across Proactive iCare and Usual Care groups using Wilcoxon tests

### Protocol Changes

The study period was decreased from 12 to 9 months to allow more subjects to be enrolled and complete the study by the mandatory study stop date. Enrollment sites were also added to the study, including the Denver Veterans Affairs Medical Center and primary care practices within the front-range urban corridor, including Denver, Colorado Springs and Pueblo. The primary outcome for the study was healthcare cost, which was to be derived from detailed records from University of Colorado and Kaiser Permanente. Because the study expanded to multiple sites and systems, it was not feasible to obtain information from the number of subjects necessary to perform a quality healthcare cost analysis, so the original primary outcome is not reported.

### Results

Five hundred and eleven subjects were enrolled into the study, including 352 in the Proactive iCare treatment group and 159 into the Usual Care control group (Fig. 1). Overall, subjects were 68 years old, predominantly male and had a pre-bronchodilator FEV_1_ of 37% predicted. Demographics and other patient characteristics at enrollment are summarized in Table 2. Usual Care and Proactive iCare groups were similar at baseline, with the exceptions of LTOT, SAMA, LAMA, ICS, COPD-related urgent office visits within the previous 12 months, and the SGRQ total and impact scores (Table 2).

**Figure 1.**
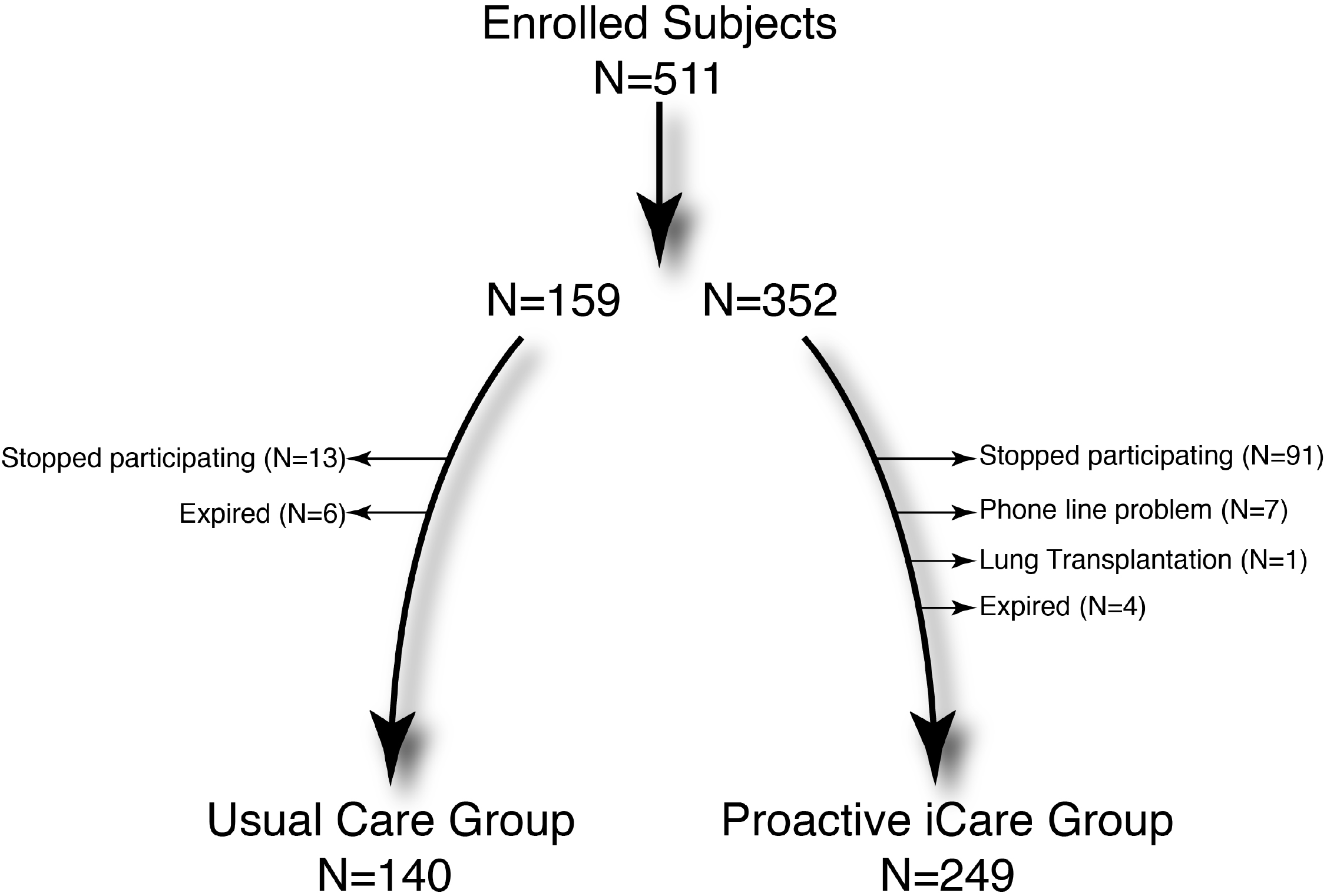
Flow Diagram. This flow diagram shows how the original study cohort (N=511) was divided between the Usual Care Group and the Proactive Integrated Care (Proactive iCare) Group. One hundred and twenty-two subjects withdrew or stopped participating in the study. The remaining subjects (N=389) were analyzed for the study outcomes.

**Table 2.**
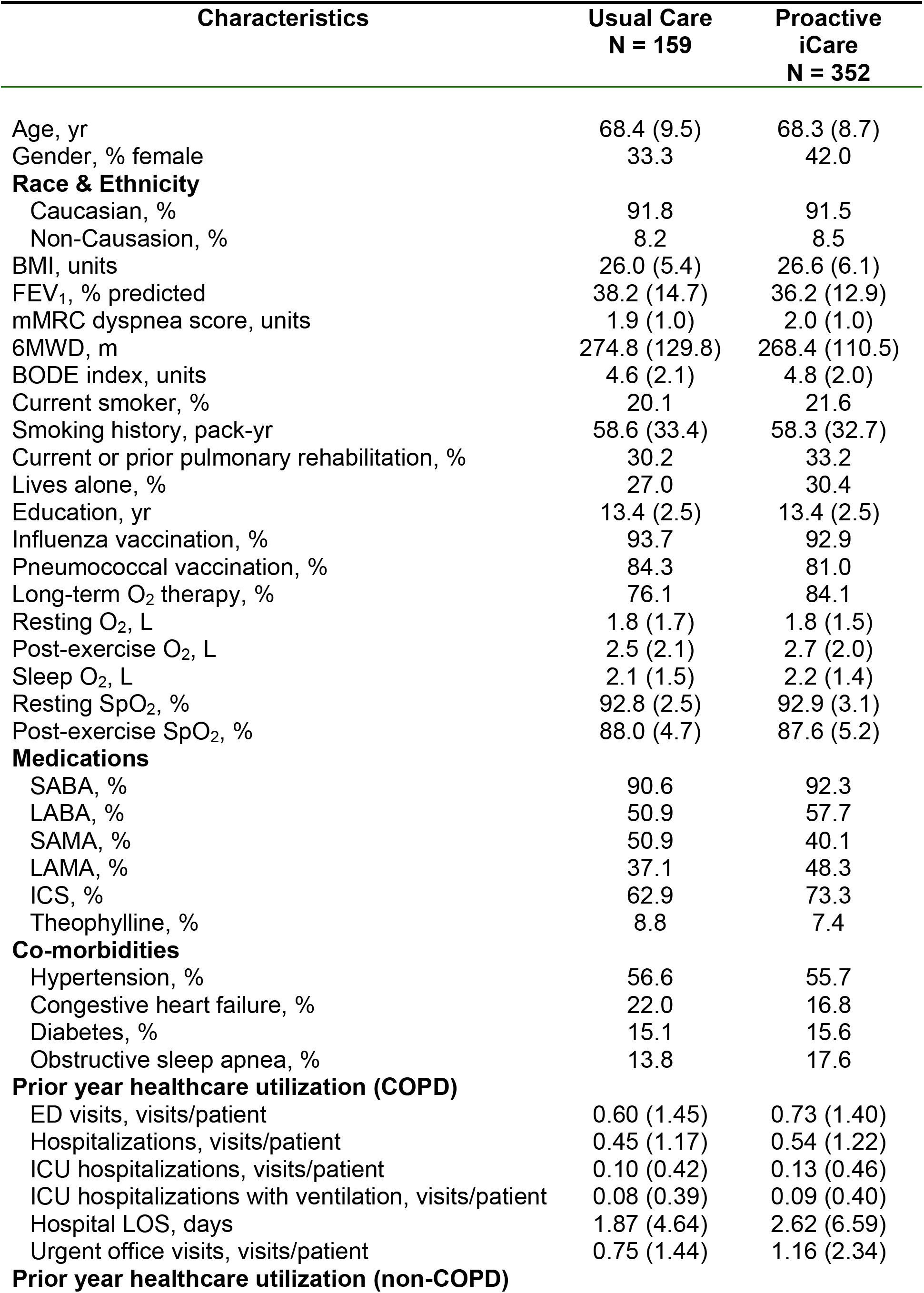
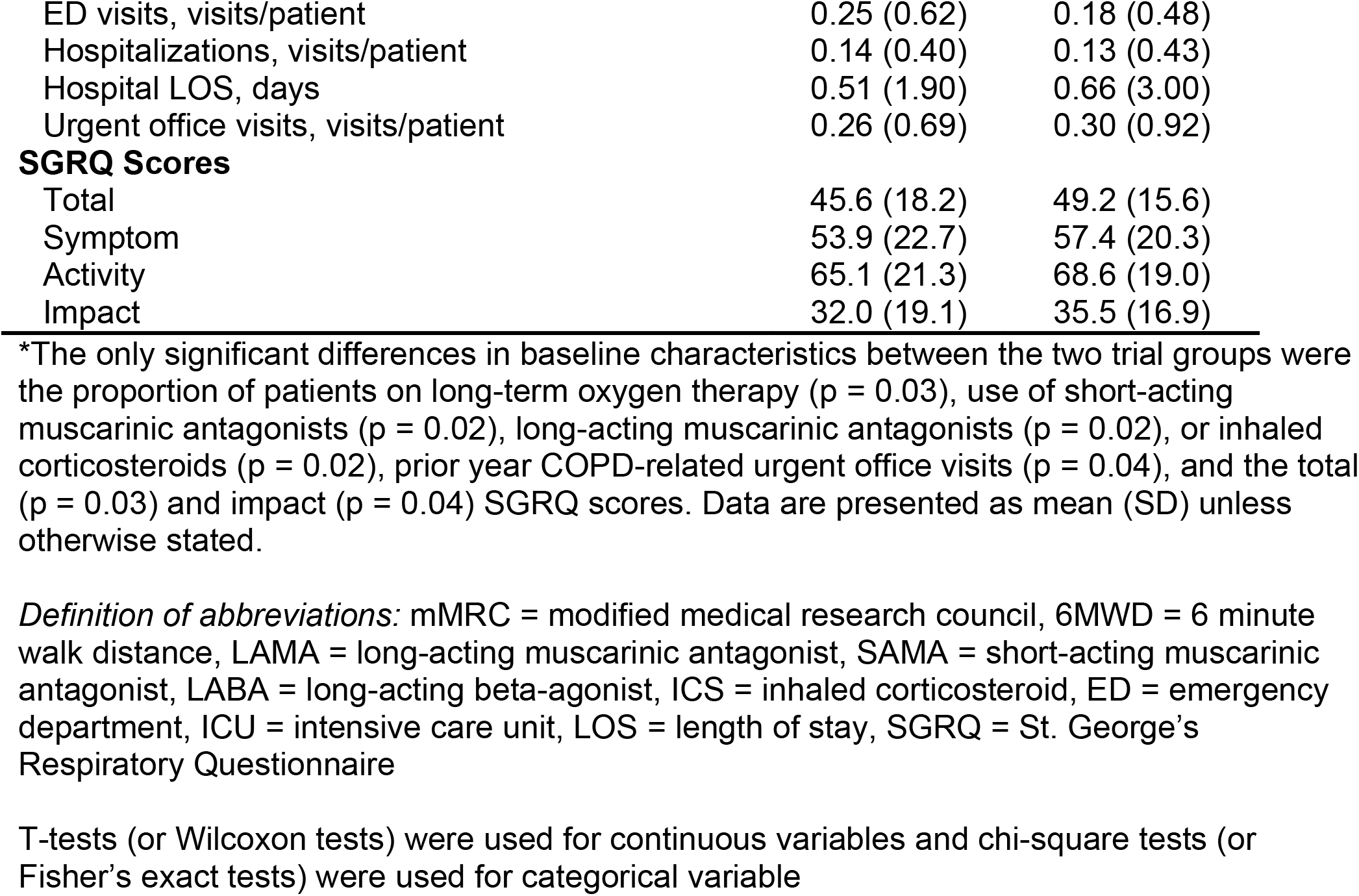
Baseline Characteristics^*^.

Twenty-four percent (N = 122) of subjects did not complete the study (Fig. 1). In the Proactive iCare group, 19.3% of subjects (N = 68) dropped out of the study within the first three months, and an additional 10% (N = 35) of subjects dropped out in the last six months of the study. In the Usual Care group, 6.9% (N = 11) subjects dropped out of the study within the first three months, and another 5% (N = 8) of subjects dropped out in the last six months. Supplementary Table 1 shows differences in baseline characteristics for completers (N = 389) and non-completers (N = 122). Non-completers were more likely to be current smokers, have a diagnosis of congestive heart failure and report more COPD-related healthcare usage within the prior year (Supplementary Table 1). Non-completers also reported less use of pulmonary rehabilitation and years of education (Supplementary Table 1). Supplementary Table 2 shows differences in baseline characteristics for subjects that completed the study. The groups remained balanced at baseline, except for differences in use of SAMA, LAMA and ICS medications, and COPD-related emergency room and urgent office visits.

### Healthcare Costs

During the course of the study enrollment was expanded to multiple sites and systems such that it was not possible to obtain the information necessary to quantify healthcare costs. Accordingly, data pertaining to the primary outcome are not reported.

### Quality of life

Patients randomized to receive Proactive iCare had large and consistent improvements (*i.e*. decreased scores exceeding the 4-point minimal clinically important difference) in the total SGRQ and SGRQ components (Table 3 and Figure 2). Using an intention-to-treat analysis, Proactive iCare improved the total SGRQ by 6.67 units, 9.46 units and 8.4 units (p < 0.0001) at 3, 6 and 9 months, respectively, compared with the Usual Care group. These improvements in the total SGRQ are similar to the results for participants with complete data in Table 3.

**Table 3.**
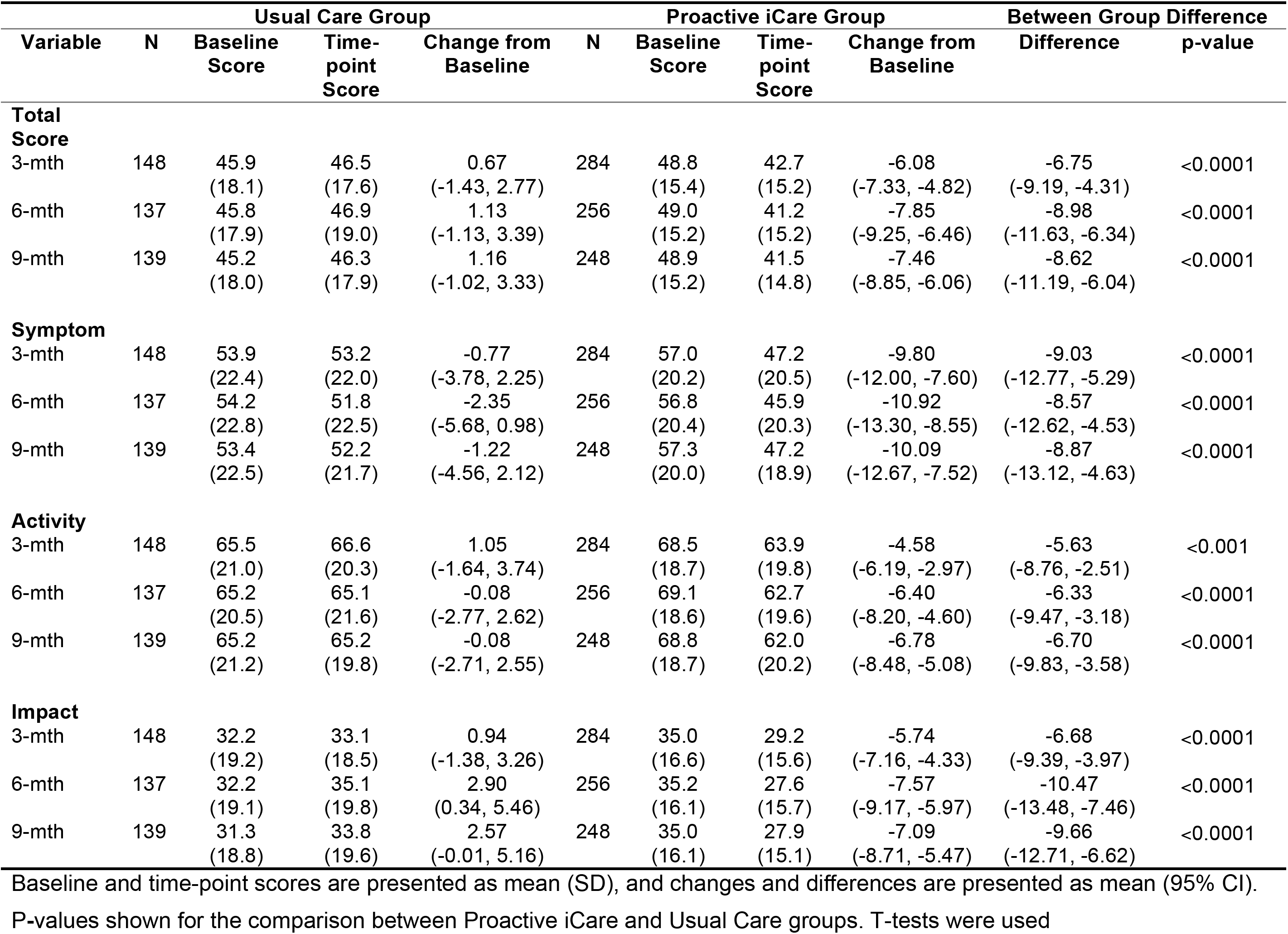
Quality of Life Measured by the St. George’s Respiratory Questionnaire.

**Figure 2.**
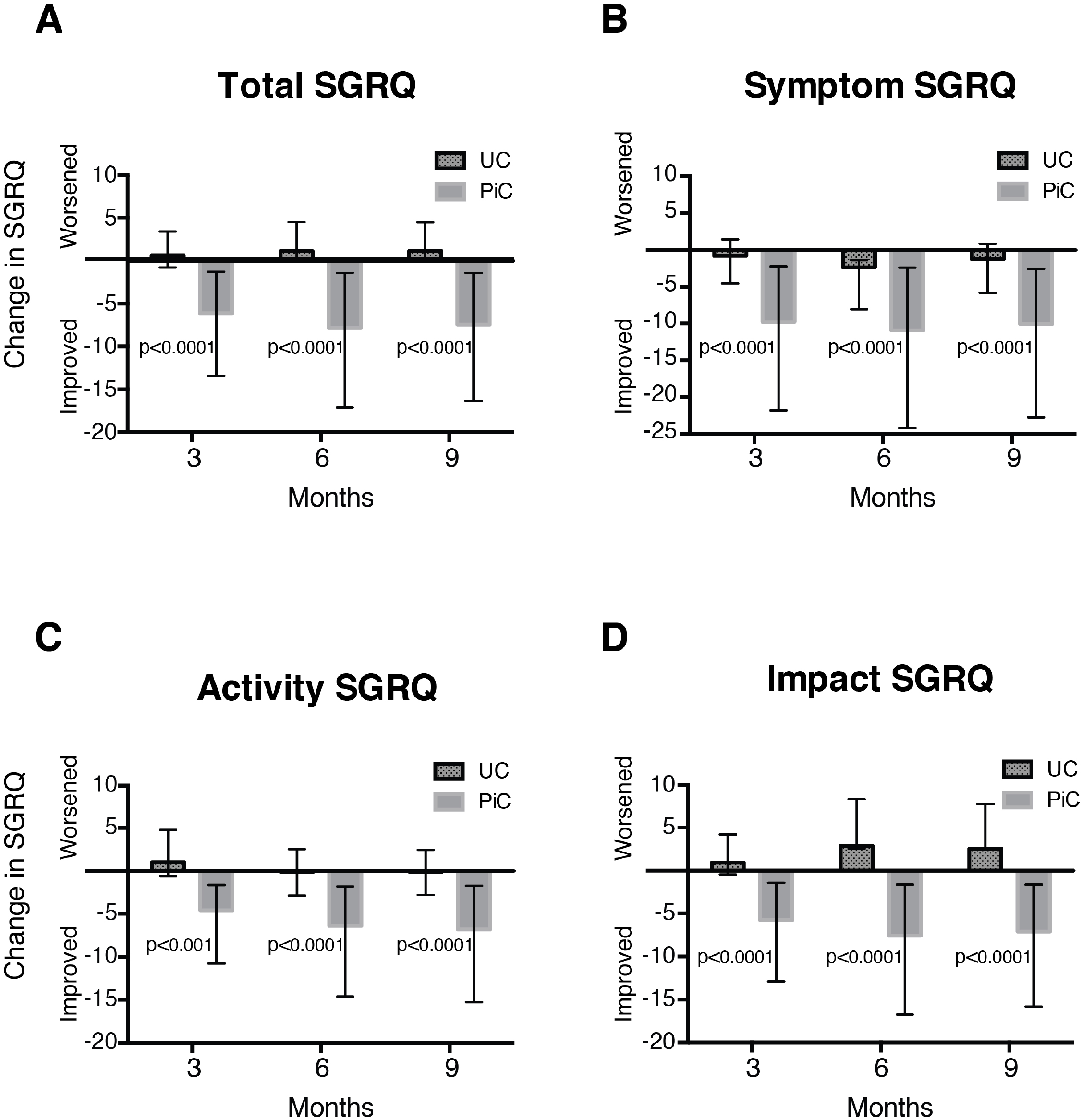
Quality of Life. This figure shows how quality of life changed from baseline at 3, 6 and 9 months in the Usual Care (UC) and Proactive Integrated Care (PiC) groups of the total cohort. Quality of life measured by the A) total, B) symptom, C) activity and D) impact domains of the St. George’s Respiratory Questionnaire (SGRQ) are shown ± 95% CI. T-tests were used to compare differences in the change from baseline for the UC and PiC groups at each time-point.

### Smoking rates, Symptoms, Exercise Capacity and Guideline-based Care

After 9 months, subjects in the Proactive iCare group reported less current smoking, cough, sputum, and breathlessness, and increased post-exercise O_2_ liter flow and SpO_2_ compared with Usual Care (Table 4). Subjects in Proactive iCare were also able to walk 42m farther (*i.e*. 15% increase from baseline) during a 6MWT, whereas those randomized to usual care did not improve (*i.e*. 1% increase). No differences were observed in the use of inhaled medications (data not shown).

**Table 4.** Smoking, Symptoms, Exercise Capacity and Guideline-based Care.

| Measurement/<br>Treatment | Usual Care |  |  | Proactive iCare |  |  | Inter-group Difference |  |
| --- | --- | --- | --- | --- | --- | --- | --- | --- |
|  | Baseline | 9 Months | Change from<br>Baseline | Baseline | 9 Months | Change from<br>Baseline | Difference<br>(95% CI) | p-value |
| FEV<sub>1</sub>, %<br>predicted | 37.5 (14.2) | 37.1 (14.6) | -0.46<br>(-2.07, 1.14) | 36.3 (13.0) | 32.9 (14.2) | -3.48<br>(-4.68, -2.28) | -3.02<br>(-5.03, -1.00) | 0.004 |
| mMRC<br>dyspnea scale | 1.9 (1.0) | 2.1 (1.0) | 0.17<br>(0.02, 0.33) | 1.9 (1.0) | 1.9 (0.9) | -0.05<br>(-0.16, 0.06) | -0.23<br>(-0.41, -0.04) | 0.02 |
| 6MWD, m | 276.8<br>(128.3) | 281.6<br>(130.4) | 2.53<br>(-15.69, 20.75) | 273.8<br>(109.1) | 318.6<br>(131.3) | 42.05<br>(29.16, 54.93) | 39.52<br>(17.50, 61.53) | 0.0005 |
| BODE index | 4.6 (2.1) | 4.7 (2.2) | 0.14<br>(-0.09, 0.37) | 4.8 (2.0) | 4.5 (2.0) | -0.21<br>(-0.41, -0.01) | -0.35<br>(-0.65, -0.05) | 0.02 |
| Current<br>smoker, % | 16.4 | 18.6 |  | 18.5 | 14.9 |  |  | 0.01 |
| Cough, % | 54.3 | 55.7 |  | 58.6 | 47.8 |  |  | 0.03 |
| Sputum, % | 44.3 | 52.9 |  | 50.2 | 44.2 |  |  | 0.007 |
| Post-exercise<br>SpO<sub>2</sub>, % | 87.9 (4.7) | 87.8 (4.2) | -0.05<br>(-0.78, 0.68) | 87.7 (5.1) | 89.4 (3.9) | 1.57<br>(0.86, 2.28) | 1.62<br>(0.60, 2.63) | 0.002 |
| Post-exercise<br>O<sub>2</sub>, L | 2.5 (2.0) | 2.4 (2.3) | -0.04<br>(-0.27, 0.18) | 2.8 (2.0) | 3.2 (2.3) | 0.42<br>(0.22, 0.61) | 0.46<br>(0.15, 0.77) | 0.004 |

Baseline and 9-month data are presented as mean (SD), and changes and differences are presented as mean (95% CI) unless otherwise stated.
*Definition of abbreviations:* mMRC = modified medical research council, 6MWD = 6 minute walk distance, LAMA = long-acting muscarinic antagonist, SAMA = short-acting muscarinic antagonist, LABA = long-acting beta-agonist, ICS = inhaled corticosteroid.
P-values shown for the comparison between Proactive iCare and Usual Care groups. T-tests were used for continuous variables, and logistic regression and chi-square tests were used for dichotomous variables

### Healthcare Utilization

Proactive iCare substantially decreased COPD-related urgent office visits by 76 visits per 100 patients (p < 0.0001), and non-significantly decreased COPD-related emergency departments visits (p = 0.09) and ICU hospitalizations (p = 0.10), compared with Usual Care (Table 5). In contrast, other measures of COPD- and non-COPD-related healthcare did not change.

**Table 5.** Healthcare Utilization Pre-post Difference in 9-month Rate, per Patient.

| Variable | Usual Care<br>N=140 | Proactive iCare<br>N=249 | Difference | p-value |
| --- | --- | --- | --- | --- |
| **COPD Healthcare** |  |  |  |  |
| Hospitalizations, visits/patient | -0.11<br>(-0.25, 0.03) | -0.21<br>(-0.36, -0.05) | -0.10<br>(-0.31, 0.11) | 0.58 |
| ICU hospitalizations, visits/patient | -0.01<br>(-0.09, 0.07) | -0.09<br>(-0.14, 0.04) | -0.08<br>(-0.17, 0.01) | 0.10 |
| ICU hospitalizations with ventilation, visits/patient | -0.02<br>(-0.09, 0.05) | -0.06<br>(-0.10, -0.03) | -0.04<br>(-0.12, 0.04) | 0.26 |
| Hospital LOS, days | -0.56*<br>(-1.53, 0.42) | -1.17<br>(-1.89, -0.44) | -0.61<br>(-1.82, 0.60) | 0.37 |
| ED visits, visits/patient | -0.18<br>(-0.34, -0.01) | -0.35<br>(-0.51, -0.19) | -0.17<br>(-0.40, 0.06) | 0.09 |
| Urgent office visits, visits/patient | 0.10<br>(-0.12, 0.32) | -0.66<br>(-0.95, -0.37) | -0.76<br>(-1.13, -0.40) | <0.0001 |
| **Non-COPD Healthcare** |  |  |  |  |
| Hospitalizations, visits/patient | 0.07<br>(-0.05, 0.19) | 0.05<br>(-0.02, 0.12) | -0.02<br>(-0.16, 0.12) | 0.91 |
| Hospital LOS, days | 0.42<br>(-0.27, 1.11) | -0.07<br>(-0.53, 0.39) | -0.49<br>(-1.29, 0.31) | 0.70 |
| ED visits, visits/patient | -0.00<br>(-0.10, 0.10) | 0.12<br>(0.01, 0.23) | 0.12<br>(-0.03, 0.27) | 0.32 |
| Urgent office visits, visits/patient | 0.11<br>(-0.02, 0.25) | 0.12<br>(-0.00, 0.325) | 0.01<br>(-0.18, 0.19) | 0.74 |

Data are presented as mean (95% CI).
*Definition of abbreviations:* ED = emergency department, ICU = intensive care unit, LOS = length of stay
\*Excluding one patient with pre-post difference = 195 days
P-values shown for the comparison between Proactive iCare and Usual Care groups. Wilcoxon tests were used.

### Mortality

During the 9-month study 4 of 352 subjects (1.1%) died in the Proactive iCare Group *versus* 6 of 159 subjects (3.8%) in the Usual Care group (p = 0.08 Fisher’s exact test).

### Subgroup Analyses

The effect of Proactive iCare was assessed in the urban (N = 403) and rural cohorts (N = 108). Baseline differences between the urban and rural cohorts are shown in Supplementary Table 3. Urban subjects reported greater participation in pulmonary rehabilitation, higher level of education, more vaccinations for influenza, and higher post-exercise oxygen liter flow, use of SABAs, SAMAs, and urgent office visits for non-COPD related problems, compared with rural subjects. In contrast, rural subjects were more likely to live alone.

Baseline characteristics in the overall urban cohort and in completers are shown in Supplementary Tables 4 and 5. Subjects in the urban cohort treated with Proactive iCare showed large improvements in the total and component SGRQ scores across all three time-points (Supplementary Table 6 and Supplementary Figure 1), compared with Usual Care. Proactive iCare was associated with significant improvements in dyspnea, sputum production, distance walked during the 6MWT, BODE index, and post exercise SpO_2_ % that mirrored results for the overall study (Supplementary Table 7). Proactive iCare decreased COPD-related urgent office visits by 77 visits per 100 subjects (p < 0.0001), and nonsignificantly decreased COPD-related emergency departments visits (p = 0.12) and hospitalizations (p = 0.053), compared with Usual Care (Supplementary Table 8). In the smaller rural cohort, we found no improvement in the SGRQ or in any component of healthcare utilization, but there were reductions in the number of current smokers (p = 0.04) patients producing sputum (0.049), subjects on long-term oxygen therapy (0 = 0.02) and post-exercise oxygen liter flow (p = 0.004; data not shown).

**Table 6.** Early Warning for COPD Exacerbations.

| **Flag Description\*** | **Flags (N)** |
| --- | --- |
| **Red Flags** |  |
| Shortness of breath at all times | 49 |
| No walk for two days or longer | 44 |
| Resting or exercise SpO<sub>2</sub> ≤ 87% | 44 |
| Trouble with daily routine | 32 |
| FEV<sub>1</sub> decline | 28 |
| Exercise SpO<sub>2</sub> ≤ 87% | 27 |
| Cough with green, red or brown sputum | 20 |
| Resting SpO<sub>2</sub> ≤ 87% | 17 |
| Fever >101.5° F | 1 |
| **Yellow Flags** |  |
| Coughing more than usual | 32 |
| Resting or exercise SpO<sub>2</sub> = 88 - 89% | 30 |
| Shortness of breath with activity | 29 |
| FEV<sub>1</sub> decrease | 29 |
| More shortness of breath than usual | 28 |
| Coughing increased sputum | 22 |
| Exercise SpO<sub>2</sub> = 88 - 89% | 21 |
| Some trouble with daily routine | 16 |
| Cough yellow sputum | 15 |
| No walk today | 14 |
| Wheezing in past 48h | 13 |
| Resting SpO<sub>2</sub> = 88 - 89% | 9 |

*Definition of abbreviations:* SpO<sub>2</sub> = pulse oximetry oxygen saturation.
\*Red and yellow flags that occurred within the three days prior to treatment for a COPD exacerbation

### Early Warning for COPD Exacerbations

Three hundred and fifty-two subjects in the Proactive iCare group had 210 COPD exacerbations during the study (*i.e*. 0.80 exacerbations/patient-yr). Two hundred and sixty-two red flags and 258 yellow flags occurred in the three days prior to an exacerbation (Table 5). The most common red and yellow flags were related to increasing shortness of breath, decreased physical activity, lower oxygen saturation and increased cough. Coordinator notes allowed for an accurate assessment of communication between subjects, coordinators and medical providers for 110 exacerbations, and treatment choice for 195 exacerbations. The first exacerbation-related phone call was made from subject to medical provider (*i.e*. selfreport) 21% of the time, from subject to coordinator (*i.e*. Proactive iCare assisted) 34% of the time, and from coordinator to subject (*i.e*. Proactive iCare identified) 45% of the time. Overall, the first exacerbation-related phone call involved a study coordinator 79% of the time. Sixty-one percent of exacerbations were treated with an oral corticosteroid and 70% were treated with an antibiotic.

### Discussion

The important findings of this study are that Proactive iCare improved quality of life, exercise capacity, identification of unreported exacerbations, and reduced urgent office visits for COPD. It may have also helped people stop smoking. These results suggest that coupling integrated care with remote monitoring may improve outcomes that are important for people with advanced COPD.

Multiple studies have evaluated the impact of integrated care on outcomes for COPD^15-21,28^. Two Cochrane reviews concluded that integrated care/self-care programs improve quality of life and exercise tolerance for COPD, and decrease hospitalizations and hospital length of stay for COPD exacerbations^18,19^. Both reviews included two important studies by Bourbeau *et al*. and Rice *et al^15,16^*. Bourbeau *et al*. randomized 191 advanced COPD patients with a recent COPD exacerbation to a self-management program or usual care, and found that self-management decreased unscheduled office visits, emergency room visits and hospitalizations for COPD^15^. Rice *et al*. randomized 743 patients with severe COPD and a recent exacerbation or oxygen use to an integrated care program or usual care^16^. They showed that integrated care decreased COPD-related hospitalizations and emergency department visits, and found that these improvements were related to the number of completed calls made by coordinators to patients^16^.

Unfortunately, not all studies of integrated care have been so positive^20,21,28^. An important example is a well-designed study by Fan *et al*., which randomized COPD patients with a prior severe exacerbation to integrated care *versus* usual care^21^. Unexpectedly, integrated care did not decrease COPD-related hospitalizations and was associated with increased mortality, causing the study to be stopped early. The reason(s) for increased mortality could not be determined, but the level of communication between coordinators and patients may be an important factor. For instance, there were 6 planned calls from coordinators to patients over a year in the Fan study *versus* 12 planned calls over the same period in the Bourbeau and Rice studies^15,16,21^. Patients were prompted to call a coordinator during an exacerbation or if symptoms worsened in all three studies. However, only 4.5% of exacerbations in the treatment group were communicated to a coordinator in the Fan study^24^, whereas 48% of exacerbations in the self-management group were communicated to a healthcare professional in the Bourbeau study^15^. Similar data were not available from the Rice study^16^. In response to a Letter to the Editor about this issue, Fan *et al*. wrote, "Future studies are needed to help understand the role of symptoms and physiologic monitoring approaches in care management to correctly identify COPD exacerbations, assess need for evaluation by [the] health care team, and initiate appropriate and prompt treatment and follow-up care”^24^. More recently, Aboumatar *et al*. randomized 240 patients hospitalized for COPD to a program that combined transition of care and long-term self-management. Contrary to expectations, treatment resulted in more COPD-related emergency department visits and hospitalizations, without an improvement in quality of life ^20^.

We found that Proactive iCare had positive effects on multiple patient-centered outcomes that replicate and extend results of past studies^25,26^. For example, two smaller studies each conducted over three months suggested that Proactive iCare could improve the SGRQ by 10-12 units^25,26^. In the current study, Proactive iCare improved the SGRQ by ~7 units at three months, which increased to ~9 units at six and nine months, compared to Usual Care. These results were consistent in both the per-protocol and intention-to-treat analyses. Therefore, the effect of Proactive iCare on quality of life appears to be large, sustained, and greater than most other studies of integrated care for COPD^15,16,18,21,28^. Proactive iCare also improved the 6MWD by 40m at nine months compared to Usual Care, which is similar to the 45m improvement that was seen at three months in our rural study^26^, and similar to other integrated care studies^18,19^. We also saw improvements in self-reported cough and shortness of breath measured by the mMRC dyspnea scale that is similar to what was found in our rural study^26^.

Proactive iCare decreased urgent COPD-related outpatient visits by 76 visits per 100 subjects. These results are similar to the Bourbeau study, which decreased unscheduled, COPD-related outpatient visits by ~60% in the self-management group^15^. Separate studies using administrative data from the Health Buddy® Consortium Medicare Demonstration Project showed similar improvements in overall and COPD-related hospitalizations and healthcare costs, especially for those who engaged with the Health Buddy®^29-31^. Many of the positive effects of Proactive iCare observed in the overall study were also present in the urban subgroup. On the other hand, fewer positive results were seen in the rural subgroup. This was surprising given the substantial effects of Proactive iCare in an earlier study of rural COPD patients^26^, and may have been due to lower enrollment (N = 83) that did not meet prespecified goals.

Proactive iCare was designed to uncover and treat exacerbations that would otherwise go unreported, or would be recognized at a later time. These goals are important because unreported exacerbations have negative effects^8-10^, and early therapy improves outcomes^32^. In past studies, Proactive iCare uncovered 47-78% of red-flagged COPD exacerbations that were not self-reported to coordinators or providers^25,26^. In the current study, coordinators initiated contact with subjects for exacerbations 45% of the time, indicating that Proactive iCare can identify and treat exacerbations that may go unreported. Problems with shortness of breath, physical activity, cough, and oxygenation were the most common problems that preceded an exacerbation. These early indicators are similar to the results of two prior studies of Proactive iCare in COPD^25,26^. For unclear reasons, significant proportions of COPD patients do not report exacerbations, hampering efforts to deliver early treatment^8-10^. Integrated/Self-care does not appear to fully resolve this problem, since 52% and 95% of exacerbations went unreported in studies by Bourbeau^15^ and Fan^21^, respectively. Ultimately, these results may facilitate development of the next generation of remote, and preferably passive, monitoring tools that will quantitatively measure early exacerbation indicators, so that exacerbations can be identified and treated earlier.

During the course of the study, over twice the number of subjects dropped out from the Proactive iCare group (*i.e*. 29%) compared to the Usual Care group (*i.e*. 11 %). This is an important issue, because engagement with the Health Buddy® improves outcomes^29-31^. The majority of subjects dropped out from the Proactive iCare group during the first three months (*i.e*. 19%), with the remainder leaving the study between the 3^rd^ and 9^th^ month (*i.e*. 10%). In contrast, dropouts in the Usual Care group were fewer and occurred more evenly over the course of the study. The 3-month dropout rate of 19% in the Proactive iCare Group was very similar to the 18% 3-month dropout rate that occurred during our rural Proactive iCare study^26^. A small proportion of subjects dropped out due to phone line problems or death. But the majority of subjects stopped participating for unknown reasons. Overall, subjects who left the study were roughly twice as likely to be current smokers, to have congestive heart failure, and to report COPD-related ED, hospital and ICU healthcare utilization within the prior year. How or whether these baseline characteristics negatively impacted the willingness or ability of subjects to remain engaged with the program is not known. But it is possible that patients with congestive heart failure may have needed different treatments, such as diuresis rather than a steroid. It may have also been difficult for patients who were frequently hospitalized to engage with the monitoring program five days per week, either because of fatigue or cognitive deficits. Future studies will need to uncover and address barriers to implementation. They should also explore passive monitoring approaches that might make it easier for sicker patients to participate, such as wearable sensors that can detect motion or oxygen saturation, sensors integrated into a mattress that can measure heart or respiratory rate, or audio sensors in smart phones or smart speakers that can identify cough or wheezing^33-35^.

This study has several limitations including the quasi-randomized design, which can result in selection bias and imbalances of measured and unmeasured confounders. Despite this weakness, there were few imbalances in baseline characteristics between the Proactive iCare and Usual Care groups. We were not able to perform a rigorous healthcare cost analysis as originally planned, because subjects came from different enrollment sites and systems. Instead, we reported the effects of Proactive iCare on pre-planned, patient-centered outcomes and healthcare utilization. Rigorous healthcare cost analyses should be performed when possible as Bourbeau *et al*. did on their self-care study^36^. Healthcare utilization was self-reported and was not confirmed by obtaining medical records. To improve accuracy, coordinators contacted patients every three months to collect these results. Other outcomes were not dependent on patient recollection. Most outcomes were not assessed using an intention-to-treat analysis because of a lack of repeated outcome measures. In contrast, changes in the total SGRQ were assessed using both intention-to-treat and per-protocol analyses although most of the drop-outs occurred within the first three months and a bias may still remain.

COPD is one of the most devastating diseases worldwide ^1,2,4^ yet recommended care for COPD remains suboptimal^5-7^. New models of care, such as integrated care, hold the potential to improve patient-centered outcomes, but so far results have been inconsistent ^16,18,19,21,28^. We hypothesized that coupling remote monitoring to integrated care would improve patient-centered outcomes and healthcare utilization by increasing delivery of guideline-based care and by acting as an early-warning system for COPD exacerbations. Our results suggest that Proactive iCare achieves these goals and does not worsen mortality. Future studies should also strive to understand barriers to implementation so that patient drop out is minimized, identify components of Proactive iCare that are most effective, and determine the optimal intervention intensity for different patient phenotypes. It will also be important to identify passive monitoring techniques and algorithms that identify exacerbations early and minimize the need for direct patient engagement. While these results are intriguing, they should be considered hypothesis generating until a randomized, controlled trial of Proactive iCare has been performed.

## Data Availability

The data referred to in the manuscript is provided in the manuscript and in the attached supplemental data.

## Acknowledgements

The authors wish to thank Jason Riley for creating and maintaining the Microsoft Access database.PBK, SJM and RWV had full access to all of the data in the study and take responsibility for the integrity of the data and the accuracy of the data analysis. PBK, RWV, AB, DPR, JMW and RWV contributed to the study design and concept. PBK, DLPD, TJF, SSJ, CK, SC, and RWV contributed to acquisition of data. PBK, SJM, DJL, AB, TJS, DPR, RKA, TMB, MG, RLK, JMW and RWV contributed to analysis and interpretation of data. PBK, SJM and RWV drafted the manuscript. PBK, SJM, DLPD, TJF, DJL, FDV, AB, TJS, DPR, RKA, TMB, CK, SC, MG, RLK, JMW and RWV contributed to critical revision of the manuscript for important intellectual content. SJM performed the statistical analysis. DLPD, PBK and RWV provided administrative, technical or material support. PBK and RWV supervised the study.

RWV, PBK, SJM, DLPD, TJF, SSJ, NFW, DJL, AB, TJS, DPR, CMK, SC, RLK and JMW received funding from the Colorado Cancer, Cardiovascular Disease and Chronic Pulmonary Disease Prevention, Early Detection and Treatment Program (CCPD). RWV and MG received funding from the National Institutes of Health. PBK worked for Robert Bosch Healthcare from 2010 to 2015 and for Philips Healthcare from 2015 to 2017, after the study was completed. RKA and TMB head no conflicts of interest.

## Abbreviations

COPD = chronic obstructive pulmonary disease; Proactive iCare = Proactive Integrated Care; SGRQ = St. Georges Respiratory Questionnaire, 6MWT = six minute walk test; mMRC Dyspnea Scale = modified Medical Research Council Dyspnea Scale; LTOT = long-term oxygen therapy; SABA = short acting beta agonist: LABA = long-acting beta agonist; SaMa = short acting muscarinic antagonist; LAMA = long-acting muscarinic antagonists; ICS= inhaled corticosteroid.

## Online Supplement

This article has an online data supplement.

